# Time-use and mental health during the COVID-19 pandemic: a panel analysis of 55,204 adults followed across 11 weeks of lockdown in the UK

**DOI:** 10.1101/2020.08.18.20177345

**Authors:** Feifei Bu, Andrew Steptoe, Hei Wan Mak, Daisy Fancourt

**Author notes:** Corresponding author: Daisy Fancourt 1-19 Torrington Place, London, UK, WC1E 7HB **Email:**. **Ethics approval and consent to participate** Ethical approval for the COVID-19 Social Study was granted by the UCL Ethics Committee. All participants provided fully informed consent. The study is GDPR compliant. **Author Contributions** DF, AS and FB conceived and designed the study. FB analyzed the data and FB, HWM and DF wrote the first draft. All authors provided critical revisions. All authors read and approved the submitted manuscript.

## Abstract

There is currently major concern about the impact of the global COVID-19 outbreak on mental health. But it remains unclear how individual behaviors could exacerbate or protect against adverse changes in mental health. This study aimed to examine the associations between specific activities (or time-use) and mental health and wellbeing amongst people during the Covid-19 pandemic. Data were from the UCL COVID-19 Social Study; a panel study collecting data weekly during the COVID-19 pandemic. The analytical sample consisted of 55,204 adults living in the UK who were followed up for the strict 11-week lockdown period from 21st March to 31st May 2020. Data were analyzed using fixed-effects and Arellano–Bond models. We found that changes in time spent on a range of activities were associated with changes in mental health and wellbeing. After controlling for bidirectionality, behaviors involving outdoor activities including gardening and exercising predicted subsequent improvements in mental health and wellbeing, while increased time spent on following news about COVID-19 predicted declines in mental health and wellbeing. These results are relevant to the formulation of guidance for people obliged to spend extended periods in isolation during health emergencies, and may help the public to maintain wellbeing during future pandemics.

## Introduction

A number of studies have demonstrated the negative psychological effects of quarantine, lockdowns and stay-at-home orders during epidemics including SARS, H1N1 influenza, Ebola, and COVID-19^1^^2^^3^^4–6^. These effects include increases in stress, anxiety, insomnia, irritability, confusion, fear and guilty^4–6^. To date, much of the research on the mental health impact of enforced isolation during the pandemic has focused on the mass behavior of “staying at home” as the catalyst for these negative psychological effects. But there has been little exploration into how specific behaviors within the home might have differentially affected mental health, either exacerbating or protecting against adverse psychological experiences.

Re-allocation of time use has been shown from other social shocks where people suddenly are forced to spend a significant amount of time at home, with individuals quickly having to adapt behaviorally to new circumstances and develop new routines. For example, during the 2008-2010 recession, adults in the US who lost their jobs reallocated 30% of their usual working time to “non-market work”, such as home production activities (e.g. cleaning, washing), childcare, DIY, shopping, and care of others, and spent 70% of the time on leisure activities, including socializing, watching television, reading, sleeping, and going out^7^. Similarly, during the COVID-19 pandemic, research suggests that while many individuals were able to continue working from home, others experienced furloughs or loss of employment, and many had to take on increased childcare responsibilities^8^. Further, individuals globally experienced a sharp curtailing of leisure activities, with shopping, day trips, going to entertainment venues, face-to-face social interactions, and most activities in public spaces prohibited. Analyses of Google Trends have suggested negative effects of these limitations on behaviors, showing a rise in search intensity for boredom and loneliness alongside searches for worry and sadness during the early weeks of lockdown in Europe and the US^9^. But it’s not yet clear what effect these changes in behaviors had on mental health.

There is a substantial literature on the relationship between the ways people spend their time and mental health. Certain behaviors have been proposed to exert protective effects on mental health. For instance, studies on leisure-time use show that taking up a hobby can have beneficial effects on alleviating depressive symptoms^10^, engaging in physical activity can reduce levels of depression and anxiety and enhance quality of life^11–14^, and broader leisure activities such as reading, listening to music, and volunteering can reduce depression and anxiety, increase personal empowerment and optimism, foster social connectedness, and improve life satisfaction^15–19^. However, other behaviors may have a negative influence on mental health. Engaging in productive activities (e.g. work, housework, caregiving) has been found in certain circumstances to be associated with higher levels of depression^20^, and sedentary screen time can increase the risk of depression^21^, especially when watching news or browsing internet relating to stressful events. This relationship between time use and mental health is bidirectional, as mental ill health has been shown to predict lower physical activity^22^, lower motivation to engage in leisure activities^23^ and increased engagement in screen time^24^.

However, there have been little data on the association between daily activities and mental health amongst people staying at home during the COVID-19 pandemic. Further, it is unclear if activities that are usually beneficial for mental health had similar psychological benefits during the pandemic. This topic is pivotal as understanding time use will help in formulating healthcare guidelines for individuals continuing to stay at home due to quarantine, shielding, or virus resurgences during the current global crisis and in potential future pandemics. Therefore, this study involved analyses of longitudinal data from over 50,000 adults captured during the first two months of ‘lockdown’ due to the COVID-19 pandemic in the UK. It explored the time-varying relationship between a wide range of activities and mental health, including productive activities, exercising, gardening, reading for pleasure, hobby, communicating with others, following news on COVID-19 and sedentary screen time.

Specifically, given research showing the inter-relationship yet conceptual distinction between different aspects of mental health, we focused on three different outcomes. Anxiety combines negative mood states with physiological hyperarousal, while depression also combines negative mood states with anhedonia (loss of pleasure), and life satisfaction is an assessment of how favorable one feels towards one’s attitude to life^25,26^. Crucially, symptoms of anxiety and depression can coexist with positive feelings of subjective wellbeing such as life satisfaction, and even in the absence of any specific symptoms of mental illness, individuals can experience low levels of wellbeing^27^. So this study sought to disentangle differential associations between time use and multiple aspects of mental health. As these relationships can be complex and are likely bidirectional, this study explored (a) concurrent changes in behaviors and mental health to identify associations over time, and (b) whether changes in behaviors temporally predicted changes in mental health, accounting for the possibility of reverse causality by using dynamic panel methods.

## Data and Methods

### Participants

Data were drawn from the UCL COVID-19 Social Study; a large panel study of the psychological and social experiences of over 50,000 adults (aged 18+) in the UK during the COVID-19 pandemic. The study commenced on 21st March 2020 involving online weekly data collection from participants for the duration of the COVID-19 pandemic in the UK. Whilst not random, the study has a well-stratified sample that was recruited using three primary approaches. First, snowballing was used, including promoting the study through existing networks and mailing lists (including large databases of adults who had previously consented to be involved in health research across the UK), print and digital media coverage, and social media. Second, more targeted recruitment was undertaken focusing on (i) individuals from a low-income background, (ii) individuals with no or few educational qualifications, and (iii) individuals who were unemployed. Third, the study was promoted via partnerships with third sector organisations to vulnerable groups, including adults with pre-existing mental illness, older adults, and carers. The study was approved by the UCL Research Ethics Committee (12467/005) and all participants gave informed consent. The full study protocol, including details on recruitment, retention, and weighting is available at www.covidsocialstudy.org

In this study, we focused on participants who had at least two repeated measures between 21st March and 31st May 2020, when the UK went into strict lockdown on the 23^rd^ March and remained largely in that situation until 1^st^ June (although the lockdown measures started to be eased earlier in different UK nations). This provided us with data from 55,204 participants (total observations 338,083, mean observations per person 6.1 range 2 to 11).

### Measures

Depression during the past week was measured using the Patient Health Questionnaire (PHQ-9); a standard instrument for diagnosing depression in primary care^28^. The questionnaire involves nine items, with responses ranging from “not at all” to “nearly every day”. Higher overall scores indicate more depressive symptoms.

Anxiety during the past week was measured using the Generalized Anxiety Disorder assessment (GAD-7); a well-validated tool used to screen and diagnose generalised anxiety disorder in clinical practice and research^29^. There are 7 items with 4-point responses ranging from “not at all” to “nearly every day”, with higher overall scores indicating more symptoms of anxiety.

Life satisfaction was measured by a single question on a scale of 0 to 10: “overall, in the past week, how satisfied have you been with your life?”

Thirteen measures of time-use/activities were considered. These included (i) working (remotely or outside of the house), (iii) volunteering, (iii) household chores (e.g. cooking, cleaning, tidying, ironing, online shopping etc.) or caring for other including friends, relatives or children, (iv) looking after children (e.g. bathing, feeding, doing homework or playing with children), (v) gardening, (vi) exercising outside (including going out for a walk or other gentle physical activity, going out for moderate or high intensity activity such as running, cycling or swimming), or inside the home or garden (e.g. doing yoga, weights or indoor exercise), (vii) reading for pleasure, (viii) engaging in home-based arts or crafts activities (e.g. painting, creative writing, sewing, playing music etc.), engaging in digital arts activities (e.g. streaming a concert, virtual tour of a museum etc.), or doing DIY, woodwork, metal work, model making or similar, (ix) communicating with family or friends (including phoning, video talking, or communicating via email, WhatsApp, text or other messaging service), (x) following-up information on COVID-19 (e.g. watching, listening, or reading news, or tweeting, blogging or posting about COVID-19), (xi) watching TV, films, Netflix etc. (NOT for information on COVID-19), (xii) listening to the radio or music, and (xiii) browsing the internet, tweeting, blogging or posting content (NOT for information on COVID-19). Each measure was coded as, rarely (< 30mins), low (30mins-2hrs) and high (>2hrs), except for low-intensity activities such as volunteering, gardening, exercising, reading, and arts/crafts. These were coded as, none, low (< 30mins) and high (>30mins). We used a ‘stylized questions’ approach where participants were asked to focus on a single day and consider how much time they spent on each activity on the list. However, given concerns about the cognitive burden of focusing on a ‘typical’ day (which involve aggregating information from multiple days and averaging), we asked participants to focus just on the last weekday (either the day before or the last day prior to the weekend if participants answered on a Saturday or Sunday). This approach follows aspects of the ‘time diary’ approach, but we chose weekday to remove variation in responses due to whether participants took part on weekends^30^.

### Analysis

Data analyses started by using standard fixed-effects (FE) models. FE analysis has the advantage of controlling for unobserved individual heterogeneity and therefore eliminating potential biases in the estimates of time-variant variables in panel data. It uses only within-individual variation, which can be used to examine how the change in time-use is related to the change in mental health within individuals over time. As individuals are compared with themselves over time, all time-invariant factors (such as gender, age, income, education, area of living etc.) are all accounted for automatically, even if unobserved. Compared with standard regression method, it allows for causal inference to be made under weaker assumptions in observational studies. However, FE analysis does not address the direction of causality. Given this limitation, we further employed the Arellano-Bond (AB) approach^31^, which uses lags of the outcome variable (and regressors) as instruments in a first-difference model (Eq. 1).

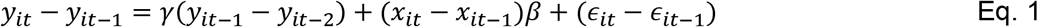

The AB model uses *y*_*it*−2_ and further lags as instruments for*y*_*it*−1_ − y_it−2_. The rationale is that the lagged outcomes are unrelated to the error term in first differences, *∊_it_* − *∊*_*it*−1_, under a testable assumption that *∊_it_* are serially uncorrelated. Further, we treated the regressors, *x_it_*, as endogenous (*E*(*x_it_∊_is_*) ≠ 0 *if s* ≤ *t, E*(*x_it_∊_is_*) = 0, *s* > *t*). Therefore, *x_it_* should be instrumented by *x*_*it*−2_, *x*_*it*−3_ and potentially further lags. The AB models were estimated using optimal generalized method of moments (GMM).

To account for the non-random nature of the sample, all data were weighted to the proportions of gender, age, ethnicity, education and country of living obtained from the Office for National Statistics^32^. To address multiple testing, we provided adjusted p values (q values) controlling for the positive false discovery rate. These were generated by using the ‘qqvalue’ package^33^. All analyses were carried out using Stata v15 and the AB models were fitted using the user-written command, xtabond2^34^.

## Results

### Descriptive

Demographic characteristics of participants are shown in Table S1 in the supplement. As shown in Table 1, the within variation accounted for about 15% of the overall variation for depression, and 16% for anxiety. Anxiety explained 56% of the variance in depression (r = 0.75, p<.001) and 27% of the variance in life satisfaction (r = –0.52, p<.001), while depression explained 32% of the variance in life satisfaction (r = –0.57, p<.001). There were also substantial changes in the time-use/activity variables (Figure 1). Over 60% of participants changed status in all activities, except for volunteering (23%) and childcare (21%).

**Figure 1.**
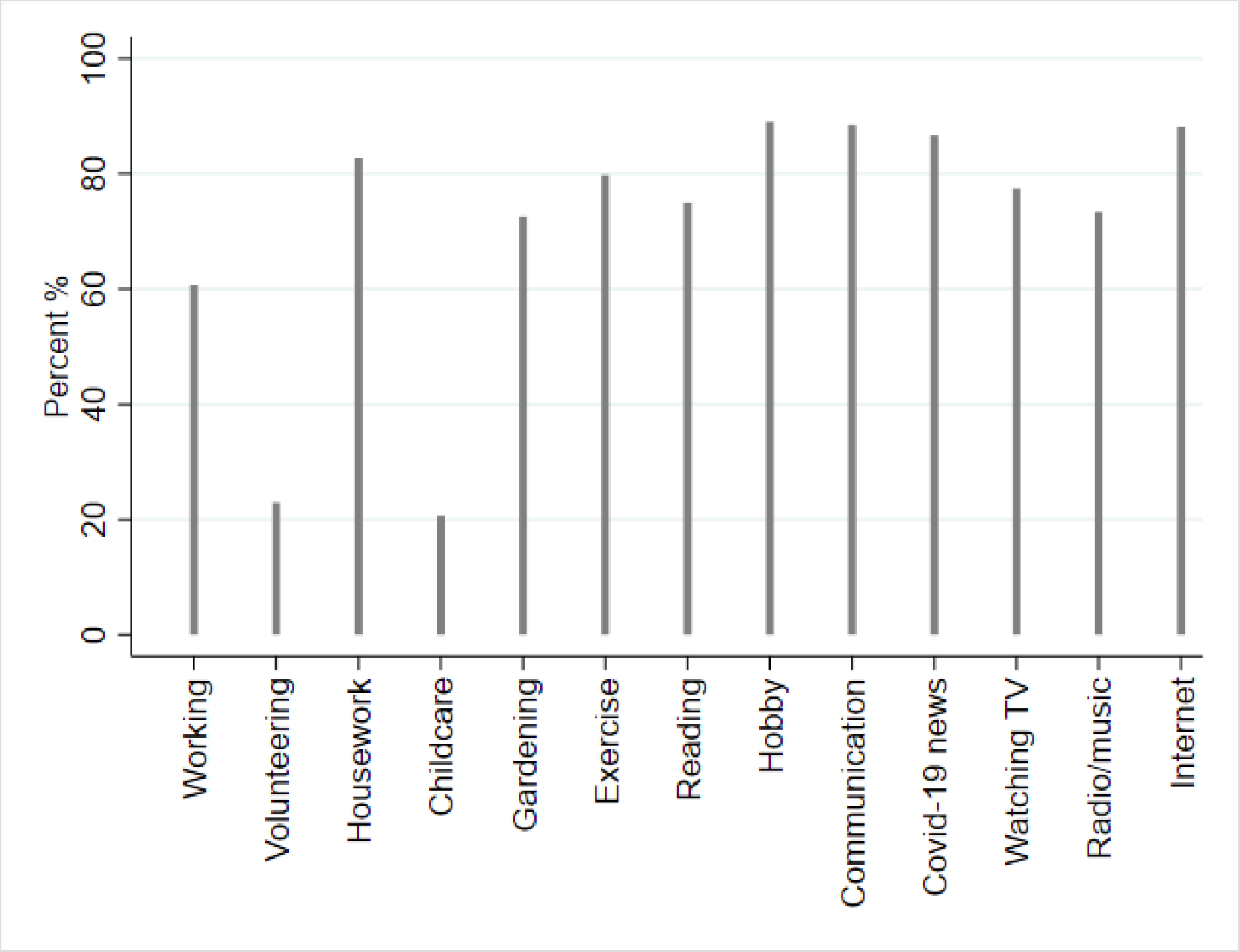
Percentages of participants changing status in the time-use/activity variables across time

**Table 1.** Summary statistics of depression, anxiety and life satisfaction Depression (PHQ-9)

|  | Depression<br>(PHQ-9) |  | Anxiety<br>(GAD-7) |  | Life<br>satisfaction |  |
| --- | --- | --- | --- | --- | --- | --- |
|  | Mean | SD | Mean | SD | Mean | SD |
| Overall | 6.09 | 5.70 | 4.73 | 5.11 | 5.89 | 2.28 |
| Between |  | 5.43 |  | 4.84 |  | 2.04 |
| Within |  | 2.22 |  | 2.08 |  | 1.10 |

### Depression

Increases in time spent working, doing housework, gardening, exercising, reading, engaging in hobbies, and listening to the radio/music were all associated with decreases in depressive symptoms (Table 2, Model I-i). The largest decrease in depression was seen for participants who increased their exercise levels to more than 30 minutes per day, who increased their time gardening to more than 30 minutes per day, or who increased their work to more than 2 hours per day. On the contrary, increasing time spent following COVID-19 news or doing other screen-based activities (either watching TV or internet use/social media) were associated with an increase in depressive symptoms.

**Table 2.** Results from the fixed-effects models on depression, anxiety and life satisfaction

|  | Model I-i<br>Depression |  |  |  | Model II-i<br>Anxiety |  |  |  | Model III-i<br>Life satisfaction |  |  |  |
| --- | --- | --- | --- | --- | --- | --- | --- | --- | --- | --- | --- | --- |
|  | Coef. | SE | p | q | Coef. | SE | p | q | Coef. | SE | p | q |
| Working 30mins-2hrs (Ref. <30mins) | -0.03 | 0.04 | 0.458 | 0.518 | 0.03 | 0.04 | 0.364 | 0.494 | 0.01 | 0.02 | 0.497 | 0.562 |
| Working >2hrs (Ref. <30mins) | **-0.27** | **0.04** | **0.000** | **0.000** | 0.06 | 0.04 | 0.095 | 0.190 | **0.11** | **0.02** | **0.000** | **0.000** |
| Volunteering <30mins (Ref. none) | 0.00 | 0.06 | 0.947 | 0.958 | -0.01 | 0.05 | 0.819 | 0.836 | 0.00 | 0.03 | 0.934 | 0.934 |
| Volunteering ≥30mins (Ref. none) | -0.16 | 0.10 | 0.104 | 0.142 | -0.09 | 0.06 | 0.148 | 0.252 | **0.09** | **0.04** | **0.028** | **0.046** |
| Housework 30mins-2hrs (Ref. <30mins) | **-0.11** | **0.03** | **0.000** | **0.000** | -0.04 | 0.03 | 0.155 | 0.252 | **0.05** | **0.01** | **0.001** | **0.002** |
| Housework >2hrs (Ref. <30mins) | **-0.21** | **0.05** | **0.000** | **0.000** | -0.04 | 0.04 | 0.360 | 0.494 | **0.06** | **0.02** | **0.004** | **0.009** |
| Looking after children 30mins-2hrs(Ref. <30mins) | -0.02 | 0.07 | 0.744 | 0.806 | 0.07 | 0.07 | 0.283 | 0.433 | 0.04 | 0.04 | 0.293 | 0.381 |
| Looking after children >2hrs (Ref. <30mins) | 0.08 | 0.10 | 0.435 | 0.514 | 0.17 | 0.09 | 0.054 | 0.128 | 0.05 | 0.05 | 0.314 | 0.389 |
| Garden <30mins (Ref. none) | **-0.15** | **0.03** | **0.000** | **0.000** | **-0.15** | **0.03** | **0.000** | **0.000** | **0.06** | **0.02** | **0.000** | **0.000** |
| Garden ≥30mins (Ref. none) | **-0.30** | **0.04** | **0.000** | **0.000** | **-0.24** | **0.03** | **0.000** | **0.000** | **0.16** | **0.02** | **0.000** | **0.000** |
| Exercising <30mins (Ref. none) | **-0.19** | **0.04** | **0.000** | **0.000** | -0.03 | 0.03 | 0.437 | 0.541 | **0.10** | **0.02** | **0.000** | **0.000** |
| Exercising ≥30mins (Ref. none) | **-0.39** | **0.04** | **0.000** | **0.000** | **-0.23** | **0.03** | **0.000** | **0.000** | **0.22** | **0.02** | **0.000** | **0.000** |
| Reading <30mins (Ref. none) | -0.07 | 0.03 | 0.041 | 0.063 | -0.06 | 0.03 | 0.048 | 0.125 | 0.03 | 0.02 | 0.090 | 0.130 |
| Reading ≥30mins (Ref. none) | **-0.14** | **0.04** | **0.001** | **0.002** | **-0.19** | **0.04** | **0.000** | **0.000** | **0.05** | **0.02** | **0.006** | **0.012** |
| Hobby <30mins (Ref. none) | -0.06 | 0.03 | 0.052 | 0.075 | -0.01 | 0.03 | 0.836 | 0.836 | 0.02 | 0.01 | 0.117 | 0.160 |
| Hobby ≥30mins (Ref. none) | **-0.17** | **0.03** | **0.000** | **0.000** | **-0.10** | **0.03** | **0.000** | **0.000** | **0.09** | **0.01** | **0.000** | **0.000** |
| Communication 30mins-2hrs (Ref. <30mins) | -0.05 | 0.03 | 0.037 | 0.060 | 0.04 | 0.03 | 0.134 | 0.249 | **0.04** | **0.01** | **0.001** | **0.002** |
| Communication >2hrs (Ref. <30mins) | 0.00 | 0.04 | 0.958 | 0.958 | **0.11** | **0.04** | **0.004** | **0.013** | **0.06** | **0.02** | **0.008** | **0.015** |
| COVID-19 news 30mins-2hrs (Ref. <30mins) | **0.29** | **0.02** | **0.000** | **0.000** | **0.48** | **0.02** | **0.000** | **0.000** | **-0.15** | **0.01** | **0.000** | **0.000** |
| COVID-19 news >2hrs (Ref. <30mins) | **0.56** | **0.05** | **0.000** | **0.000** | **0.89** | **0.04** | **0.000** | **0.000** | **-0.28** | **0.02** | **0.000** | **0.000** |
| Watching TV 30mins-2hrs (Ref. <30mins) | -0.03 | 0.04 | 0.423 | 0.514 | -0.03 | 0.04 | 0.380 | 0.494 | 0.02 | 0.02 | 0.332 | 0.392 |
| Watching TV >2hrs (Ref. <30mins) | **0.13** | **0.05** | **0.016** | **0.028** | 0.02 | 0.05 | 0.638 | 0.721 | -0.04 | 0.02 | 0.065 | 0.099 |
| Listening radio/music 30 mins-2hrs (Ref. <30mins) | **-0.09** | **0.03** | **0.003** | **0.006** | -0.04 | 0.03 | 0.089 | 0.190 | **0.03** | **0.02** | **0.027** | **0.046** |
| Listening radio/music >2hrs (Ref. <30mins) | **-0.24** | **0.05** | **0.000** | **0.000** | -0.09 | 0.04 | 0.031 | 0.090 | **0.09** | **0.02** | **0.000** | **0.000** |
| Internet/social media 30mins-2hrs (Ref. <30mins) | 0.04 | 0.03 | 0.160 | 0.208 | -0.01 | 0.02 | 0.775 | 0.836 | -0.01 | 0.01 | 0.701 | 0.759 |
| Internet/social media high >2hrs (Ref. <30mins) | **0.11** | **0.04** | **0.015** | **0.028** | 0.02 | 0.04 | 0.519 | 0.613 | 0.00 | 0.02 | 0.878 | 0.913 |
| Number of observations | 308,182 |  |  |  | 308,182 |  |  |  | 308,182 |  |  |  |
| Number of individuals | 54,632 |  |  |  | 54,632 |  |  |  | 54,632 |  |  |  |

When examining the direction of the relationship (Table 3, Model I-ii), increases in gardening, exercising, reading, and listening to the radio/music predicted subsequent decreases in depressive symptoms. However, increases in time spent following news on COVID-19 predicted increases in depressive symptoms, as did increases in time spent looking after children or moderate increases in communicating via videos, calling or messaging with others.

**Table 3.**
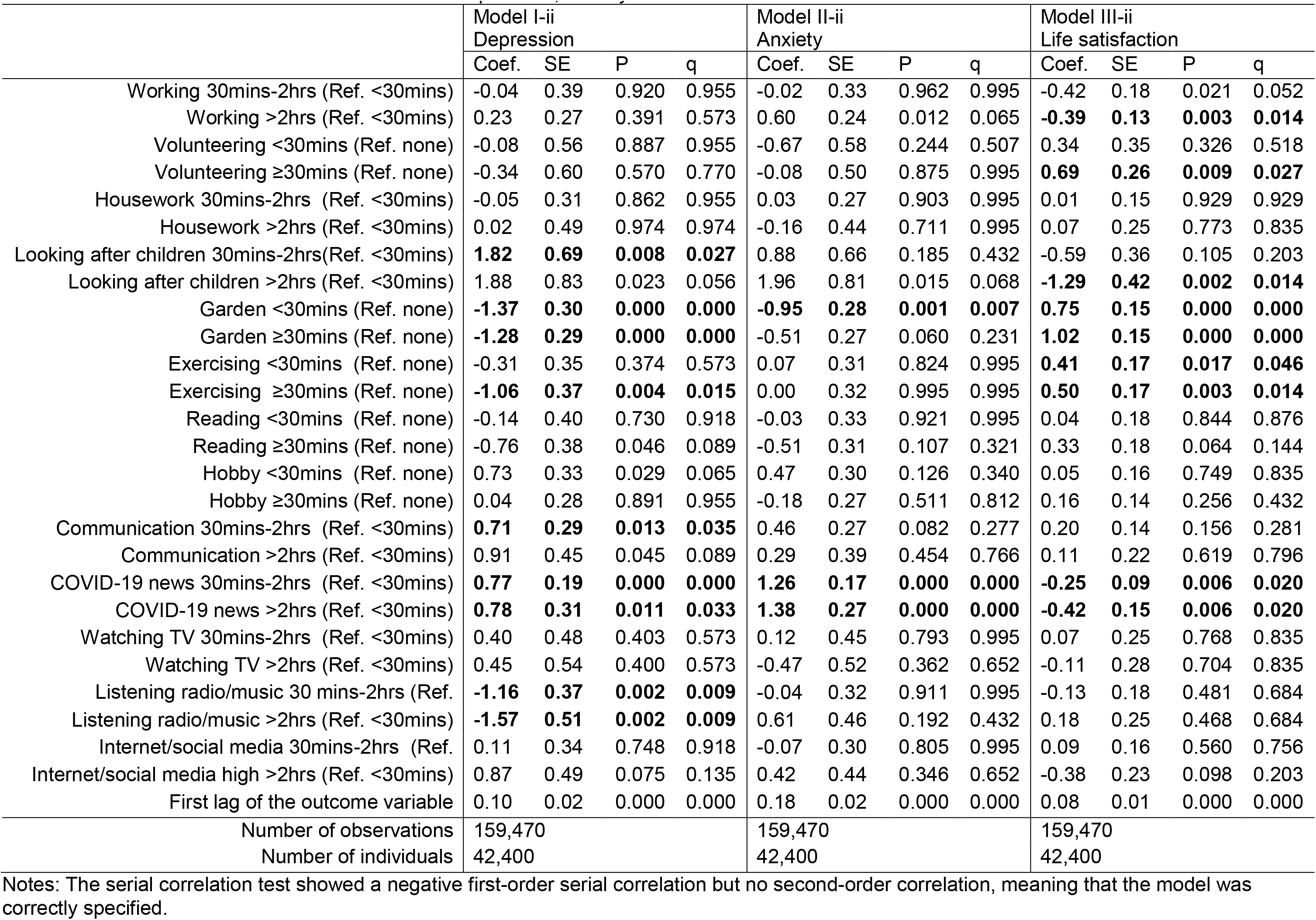
Results from the Arellano-Bond models on depression, anxiety and life satisfaction

### Anxiety

Increases in time spent gardening, exercising, reading and other hobbies were all associated with decreases in anxiety, while increasing time spent following COVID-19 news and communicating remotely with family/friends were associated with increases in anxiety (Table 2, Model II-i). The largest decrease in anxiety was seen for participants who increased their time on gardening, exercising or reading to 30 minutes or more per day.

When looking at the direction of the relationship (Table 3, Model II-ii), increases in gardening predicted a subsequent decrease in symptoms of anxiety. But increasing time spent following news on COVID-19 predicted an increase in anxiety.

### Life satisfaction

Increases in time spent working, volunteering, doing housework, gardening, exercising, reading, engaging in hobbies, communicating remotely with family/friends, and listening to the radio/music were all associated with an increase in life satisfaction, while increasing time spent following COVID-19 news was associated with a decrease in life satisfaction (Table 2, Model III-i).

When looking at the direction of the relationship (Table 3, Model III-ii), increases in volunteering, gardening and exercising predicted a subsequent increase in life satisfaction. But increasing time spent following news on COVID-19, working, and looking after children predicted a decrease in life satisfaction.

### Sensitivity analyses

We carried out sensitivity analyses excluding keyworkers who might not have been isolated at home in the same way and therefore might have had different patterns of behaviors during lockdown. The results were materially consistent with the main analysis (See the Supplementary Material).

## Discussion

This is the first study to examine the impact of time-use on mental health amongst people during the COVID-19 pandemic. Time spent on work, housework, gardening, exercising, reading, hobbies, communicating with friends/family, and listening to music were all associated with improvements in mental health and wellbeing, while following the news on COVID-19 (even for only half an hour a day) and watching television excessively were associated with declines in mental health and wellbeing. Whilst the relationship between time use and behaviors is bidirectional, when exploring the direction of the relationship using lagged models, behaviors involving outdoor activities including gardening and exercising predicted subsequent improvements in mental health and wellbeing, while time spent watching the news about COVID-19 predicted declines in mental health and wellbeing.

Our findings of negative associations between following the news on COVID-19 and mental health echo a cross-sectional study from China showing that social media exposure during the pandemic is associated with depression and anxiety^1^. The fact that exposure to COVID-19 news is largely screen-based, and the fact that watching high levels of television or high social media engagement unrelated to COVID-19 was also found to be associated with depression could suggest that this finding is more about the screens than the news specifically^35^. However, the association with following the news on COVID-19 was independent of these other screen behaviors and was found for even relatively low levels of exposure (30mins-2 hours). Further, there have been wider discussions of the negative impact of news during the pandemic, including concerns about the proliferation of misinformation and sensationalised stories on social media^36^, and information overload, whereby the amount of information exceeds people’s ability to process^37^. It is notable that these associations were found for all measures of mental ill-health and wellbeing and even in lagged models that attempted to remove the effects of reverse causality, suggesting the strength of its relationship with mental health.

However, other activities were shown to have protective associations with mental health. In particular, outdoor activities such as gardening and exercise were associated with better levels of mental health and wellbeing across all measures, with many of these results maintained in lagged models. These results echo many previous studies into the benefits of outdoors activities^10–13^. Exercise (including gentle activities such as gardening) can affect mental health via physiological mechanisms (such as reducing blood pressure), neuroendocrine mechanisms (such as reducing levels of cortisol involved in stress response), neuroimmune mechanisms (including reducing levels of inflammation associated with depressive symptoms and increasing the synthesis and release of neurotransmitters and neurotrophic factors associated with neurogenesis and neuroplasticity), and psychological mechanisms (including improving self-esteem, autonomy and mood)^38^. Particularly during lockdown, such activities (which provided opportunities to leave the home) may have helped in providing physical and mental separation from fatiguing or stressful situations at home, offering a change of scenery, and proving a feeling of being connected to something larger^39^.

Hobbies such as listening to music, reading, and engaging in arts and other projects were also associated with better mental health across all measures. This builds on substantial literature showing the benefits of such activities in reducing depression and anxiety, building a sense of self-worth and self-esteem, fostering self-empowerment, and supporting resilience^16^. The associations presented here show that these activities have remained beneficial to mental health during lockdown. However, these associations were not retained as consistently across lagged models. This suggests that they may be linked more bidirectionally with mental health, with changes in mental health also driving individuals’ motivations to engage with these activities.

There are several other noteworthy findings from these analyses. First, volunteering was associated with higher levels of life satisfaction, including across lagged models that explored with the direction of association, but not with other aspects of mental health. Previous studies have suggested psychological benefits of volunteering, but our findings suggest that it plays a specific role in supporting evaluative wellbeing during the pandemic^17^,^19^. Second, both work and housework had some protective associations when looking at parallel changes with mental health over time. However, when looking at lagged models, housework does not appear to have been a precursor to changes in mental health, whilst frequent working was associated with lower life satisfaction, independent of other types of predictors. This echoes research highlighting working from home as a cause of stress for many people during the COVID-19 pandemic^8^. Similarly, looking after children was not associated with changes in mental health in our main models, but increases to high volumes of childcare were associated with higher levels of depression and lower life satisfaction over time. This could reflect strain from spending substantial amounts of time on childcare or, as such increases may reflect changes in other aspects of home life such as a partner having to reduce childcare to go back to work, it could also reflect other stressors that may have in fact been driving changes in mental health. Finally, communicating with family/friends had mixed effects in our main models, but when exploring the direction of association, it was in fact associated with higher levels of depression. This could be explained by data from previous studies showing that while face-to-face interactions can decrease loneliness (which is associated with mental health including depression), communication over the telephone (or other digital means) can in certain circumstances increase loneliness, perhaps as it is perceived as a less emotionally rewarding experience^40^.

This study has a number of strengths including its large sample size, repeated weekly follow-up over the 11 weeks of UK lockdown, and robust statistical approaches being applied. However, the UCL COVID-19 Social Study did not use a random sample. Nevertheless, the study does have a large sample size with wide heterogeneity, including good stratification across all major socio-demographic groups, and analyses were weighted on the basis of population estimates of core demographics, with the weighted data showing good alignment with national population statistics and another large scale nationally representative social survey. But we cannot rule out the possibility that the study inadvertently attracted individuals experiencing more extreme psychological experiences, with subsequent weighting for demographic factors failing to fully compensate for these differences. This study looked at adults in the UK in general, but it is likely that “lock-down” or “stay at home” orders had different impact on time-use for people with different socio-demographic characteristics, for example age and gender. While our analyses statistically took account of all stable participant characteristics (even if unobserved) by comparing participants against themselves, future studies could examine how the relationship between time-use and mental health differs by individuals’ characteristics and backgrounds. We also lack data to see how behaviors during lockdown compared to behaviors prior to COVID-19, so it remains unknown whether changes such as increasing time spent on childcare or leisure activities were unusual for participants and therefore not part of their usual coping strategies for their mental health. Finally, we asked individuals to focus on the last available weekday in answering the questions on time use. Whilst this has been shown to improve the quality and accuracy of recollection, it does mean that variations in time use across the entire week are not captured. Finally, whilst we standardised our questions to the last week day and used the same response with all participants consistently across lockdown (which is well recognised as an approach in tracking time use, as discussed in the Methods section), it is nevertheless possible that behaviors across weekends may also have been influencing mental health independent of weekday behaviors.

Overall, our analyses provide the first comprehensive exploration of the relationship between time-use and mental health during lockdowns due to the COVID-19 pandemic. Many behaviors commonly identified as important for good mental health such as hobbies, listening to music, and reading for pleasure were found to be associated with lower symptoms of mental illness and higher wellbeing. These results were seen when exploring parallel changes in time use and behaviors, attesting to the importance of both encouraging health-promoting behaviors to support mental health, and understanding mental health when setting guidelines on healthy behaviors during a pandemic. We also explored the direction of the relationship, finding that changes in outdoor activities including exercise and gardening were strongly associated with subsequent changes in mental health. However, increasing exposure to news on COVID-19 was strongly associated with declines in mental health. These results are important in formulating guidance for people likely to experience enforced isolation for months to come (either due to quarantine, self-isolation or shielding) and are also key in preparing for future pandemics so that more targeted advice can be given to individuals to help them stay well at home.

## Data Availability

The data will be made available once the survey has been completed.

## Supplementary Material

**Table S1.** Demographic characteristics (N = 55,204 weighted)

|  | Variables | Percentages |
| --- | --- | --- |
| Age | 18-29 | 18.4% |
|  | 30-45 | 27.3% |
|  | 46-59 | 24.4% |
|  | 60+ | 29.9% |
| Gender | Women (vs. men) | 50.0% |
| Ethnicity | BAME (vs. white) | 12.5% |
| Education | GCSE or below | 32.0% |
|  | A-levels or equivalent | 33.5% |
|  | Degree or above | 34.5% |
| Household income | Low (<30k) (vs. high) | 47.3% |
| Area of living | City | 34.4% |
|  | Large town | 20.7% |
|  | Small town | 24.6% |
|  | Village, hamlet or other | 20.3% |
| Key worker role | Key worker | 22.5% |

*Note: all observed and unobserved time-invariant demographic characteristics are automatically accounted for in fixed-effects models but are shown here for descriptive purposes.*

**Table S2.** Results from the fixed-effects models on depression, anxiety, life satisfaction and happiness (excluding key workers)

|  | Model I-i<br>Depression |  |  | Model II-i<br>Anxiety |  |  | Model III-i<br>Life satisfaction |  |  |
| --- | --- | --- | --- | --- | --- | --- | --- | --- | --- |
|  | Coef. | SE | P | Coef. | SE | P | Coef. | SE | P |
| Working 30mins-2hrs (Ref. <30mins) | 0.00 | 0.05 | 0.977 | 0.04 | 0.04 | 0.368 | 0.01 | 0.02 | 0.598 |
| Working >2hrs (Ref. <30mins) | -0.24 | 0.05 | 0.000 | 0.09 | 0.04 | 0.041 | 0.09 | 0.02 | 0.000 |
| Volunteering <30mins (Ref. none) | 0.00 | 0.07 | 0.999 | -0.06 | 0.05 | 0.295 | 0.03 | 0.03 | 0.278 |
| Volunteering ≥30mins (Ref. none) | -0.23 | 0.11 | 0.046 | -0.12 | 0.07 | 0.093 | 0.12 | 0.04 | 0.009 |
| Housework 30mins-2hrs (Ref. <30mins) | -0.10 | 0.03 | 0.005 | -0.02 | 0.03 | 0.427 | 0.03 | 0.01 | 0.018 |
| Housework >2hrs (Ref. <30mins) | -0.20 | 0.05 | 0.000 | -0.01 | 0.05 | 0.785 | 0.04 | 0.02 | 0.132 |
| Looking after children 30mins-2hrs(Ref. <30mins) | -0.01 | 0.08 | 0.888 | 0.04 | 0.08 | 0.669 | 0.05 | 0.04 | 0.253 |
| Looking after children >2hrs (Ref. <30mins) | 0.08 | 0.11 | 0.466 | 0.19 | 0.11 | 0.080 | 0.05 | 0.06 | 0.422 |
| Garden <30mins (Ref. none) | -0.16 | 0.03 | 0.000 | -0.14 | 0.03 | 0.000 | 0.05 | 0.02 | 0.004 |
| Garden ≥30mins (Ref. none) | -0.31 | 0.04 | 0.000 | -0.23 | 0.04 | 0.000 | 0.16 | 0.02 | 0.000 |
| Exercising <30mins (Ref. none) | -0.14 | 0.04 | 0.000 | 0.00 | 0.04 | 0.921 | 0.09 | 0.02 | 0.000 |
| Exercising ≥30mins (Ref. none) | -0.35 | 0.04 | 0.000 | -0.19 | 0.04 | 0.000 | 0.21 | 0.02 | 0.000 |
| Reading <30mins (Ref. none) | -0.04 | 0.04 | 0.317 | -0.02 | 0.03 | 0.520 | 0.03 | 0.02 | 0.120 |
| Reading ≥30mins (Ref. none) | -0.12 | 0.05 | 0.013 | -0.16 | 0.04 | 0.000 | 0.06 | 0.02 | 0.005 |
| Hobby <30mins (Ref. none) | -0.04 | 0.03 | 0.210 | 0.01 | 0.03 | 0.683 | 0.03 | 0.02 | 0.054 |
| Hobby ≥30mins (Ref. none) | -0.17 | 0.03 | 0.000 | -0.08 | 0.03 | 0.003 | 0.11 | 0.02 | 0.000 |
| Communication low 30mins-2hrs (Ref. <30mins) | -0.08 | 0.03 | 0.008 | 0.02 | 0.03 | 0.547 | 0.04 | 0.01 | 0.002 |
| Communication high >2hrs (Ref. <30mins) | -0.01 | 0.05 | 0.882 | 0.11 | 0.04 | 0.013 | 0.06 | 0.02 | 0.010 |
| Covid-19 news 30mins-2hrs (Ref. <30mins) | 0.27 | 0.03 | 0.000 | 0.45 | 0.03 | 0.000 | -0.12 | 0.01 | 0.000 |
| Covid-19 news >2hrs (Ref. <30mins) | 0.52 | 0.05 | 0.000 | 0.84 | 0.04 | 0.000 | -0.25 | 0.02 | 0.000 |
| Watching TV 30mins-2hrs (Ref. <30mins) | -0.02 | 0.05 | 0.644 | -0.04 | 0.05 | 0.380 | 0.01 | 0.02 | 0.680 |
| Watching TV >2hrs (Ref. <30mins) | 0.13 | 0.06 | 0.029 | 0.02 | 0.06 | 0.700 | -0.05 | 0.03 | 0.063 |
| Listening radio/music 30 mins-2hrs (Ref. <30mins) | -0.09 | 0.04 | 0.012 | -0.02 | 0.03 | 0.477 | 0.03 | 0.02 | 0.082 |
| Listening radio/music >2hrs (Ref. <30mins) | -0.25 | 0.06 | 0.000 | -0.11 | 0.05 | 0.020 | 0.08 | 0.03 | 0.002 |
| Internet/social media 30mins-2hrs (Ref. <30mins) | 0.02 | 0.03 | 0.422 | -0.03 | 0.03 | 0.237 | -0.01 | 0.01 | 0.723 |
| Internet/social media high >2hrs (Ref. <30mins) | 0.06 | 0.05 | 0.242 | -0.04 | 0.04 | 0.363 | 0.01 | 0.02 | 0.830 |
| Number of observations | 239,005 |  |  | 239,005 |  |  | 239,005 |  |  |
| Number of individuals | 41,728 |  |  | 41,728 |  |  | 41,728 |  |  |

**Table S3.** Results from the Arellano-Bond models on depression, anxiety, life satisfaction and happiness (excluding key workers)

|  | Model I-ii<br>Depression |  |  | Model II-ii<br>Anxiety |  |  | Model III-ii<br>Life satisfaction |  |  |
| --- | --- | --- | --- | --- | --- | --- | --- | --- | --- |
|  | Coef. | SE | P | Coef. | SE | P | Coef. | SE | P |
| Working 30mins-2hrs (Ref. <30mins) | 0.15 | 0.38 | 0.685 | 0.29 | 0.35 | 0.398 | -0.48 | 0.18 | 0.007 |
| Working >2hrs (Ref. <30mins) | -0.04 | 0.33 | 0.913 | 0.96 | 0.29 | 0.001 | -0.30 | 0.15 | 0.047 |
| Volunteering <30mins (Ref. none) | -0.05 | 0.58 | 0.937 | -0.54 | 0.61 | 0.380 | 0.15 | 0.37 | 0.679 |
| Volunteering ≥30mins (Ref. none) | -0.03 | 0.73 | 0.971 | -0.09 | 0.53 | 0.873 | 0.34 | 0.27 | 0.207 |
| Housework 30mins-2hrs (Ref. <30mins) | 0.03 | 0.30 | 0.934 | 0.04 | 0.28 | 0.894 | -0.05 | 0.15 | 0.736 |
| Housework >2hrs (Ref. <30mins) | -0.03 | 0.49 | 0.948 | -0.44 | 0.44 | 0.308 | 0.07 | 0.25 | 0.776 |
| Looking after children 30mins-2hrs(Ref. <30mins) | 1.43 | 0.79 | 0.069 | 1.66 | 0.80 | 0.037 | -0.63 | 0.39 | 0.108 |
| Looking after children >2hrs (Ref. <30mins) | 0.91 | 0.93 | 0.332 | 2.11 | 0.95 | 0.026 | -1.36 | 0.46 | 0.003 |
| Garden <30mins (Ref. none) | -1.42 | 0.32 | 0.000 | -1.02 | 0.29 | 0.000 | 0.79 | 0.16 | 0.000 |
| Garden ≥30mins (Ref. none) | -1.43 | 0.30 | 0.000 | -0.53 | 0.27 | 0.049 | 1.10 | 0.16 | 0.000 |
| Exercising <30mins (Ref. none) | -0.41 | 0.37 | 0.265 | 0.28 | 0.32 | 0.380 | 0.33 | 0.19 | 0.087 |
| Exercising ≥30mins (Ref. none) | -0.96 | 0.39 | 0.015 | 0.30 | 0.33 | 0.369 | 0.49 | 0.18 | 0.007 |
| Reading <30mins (Ref. none) | -0.33 | 0.38 | 0.379 | -0.08 | 0.33 | 0.812 | 0.30 | 0.19 | 0.105 |
| Reading ≥30mins (Ref. none) | -1.06 | 0.37 | 0.005 | -0.52 | 0.33 | 0.116 | 0.46 | 0.18 | 0.012 |
| Hobby <30mins (Ref. none) | 1.04 | 0.35 | 0.003 | 0.67 | 0.31 | 0.033 | -0.07 | 0.16 | 0.667 |
| Hobby ≥30mins (Ref. none) | -0.02 | 0.30 | 0.938 | 0.13 | 0.29 | 0.650 | 0.09 | 0.15 | 0.542 |
| Communication low 30mins-2hrs (Ref. <30mins) | 0.32 | 0.30 | 0.292 | 0.05 | 0.27 | 0.844 | 0.28 | 0.15 | 0.058 |
| Communication high >2hrs (Ref. <30mins) | 0.62 | 0.47 | 0.186 | -0.40 | 0.41 | 0.330 | 0.20 | 0.24 | 0.401 |
| Covid-19 news 30mins-2hrs (Ref. <30mins) | 0.84 | 0.19 | 0.000 | 1.20 | 0.17 | 0.000 | -0.17 | 0.09 | 0.069 |
| Covid-19 news >2hrs (Ref. <30mins) | 0.70 | 0.33 | 0.033 | 1.56 | 0.28 | 0.000 | -0.26 | 0.16 | 0.102 |
| Watching TV 30mins-2hrs (Ref. <30mins) | -0.08 | 0.48 | 0.865 | 0.24 | 0.49 | 0.633 | -0.01 | 0.25 | 0.965 |
| Watching TV >2hrs (Ref. <30mins) | 0.02 | 0.55 | 0.967 | -0.32 | 0.54 | 0.557 | -0.28 | 0.28 | 0.325 |
| Listening radio/music 30 mins-2hrs (Ref. <30mins) | -0.72 | 0.37 | 0.051 | -0.01 | 0.32 | 0.974 | -0.05 | 0.18 | 0.777 |
| Listening radio/music >2hrs (Ref. <30mins) | -1.22 | 0.52 | 0.020 | 0.90 | 0.47 | 0.054 | 0.20 | 0.24 | 0.410 |
| Internet/social media 30mins-2hrs (Ref. <30mins) | 0.53 | 0.33 | 0.107 | 0.26 | 0.28 | 0.351 | -0.04 | 0.16 | 0.820 |
| Internet/social media high >2hrs (Ref. <30mins) | 0.91 | 0.50 | 0.066 | 0.38 | 0.45 | 0.394 | -0.25 | 0.24 | 0.295 |
| First lag of the outcome variable | 0.09 | 0.02 | 0.000 | 0.17 | 0.02 | 0.000 | 0.09 | 0.01 | 0.000 |
| Number of observations | 125,211 |  |  | 125,211 |  |  | 125,211 |  |  |
| Number of individuals | 32,855 |  |  | 32,855 |  |  | 32,855 |  |  |

